# Antimicrobial resistance in Ecuador: A One Health situational analysis of governance, infrastructure, and equity gaps

**DOI:** 10.1101/2025.09.24.25336602

**Authors:** Richar Rodriguez-Hidalgo, Linette Jácome, Diana Paz, Arne Ruckert, Raphael Aguiar, Mayumi Duarte Wakimoto, Phaedra Henley, Mary Wiktorowicz

## Abstract

Antimicrobial resistance (AMR) represents a growing global public health threat, undermining therapeutic efficacy and disproportionately affecting vulnerable populations. In Ecuador, the situation is exacerbated by weak surveillance systems and fragmentation among the human, animal, and environmental health sectors. This study analyzes current national approaches that address AMR in Ecuador from a One Health (OH) perspective, with an emphasis on governance, public policy, health infrastructure, and equity. A qualitative approach was employed combining document review, scientific literature analysis, and semi-structured interviews with key informants engaged across all OH sectors. Findings reveal structural barriers, including lack of institutionalization of the OH strategy, limited funding for the health system, weak intersectoral coordination, low community awareness, and the absence of equity-oriented policies. As a conclusion, the results highlight the urgent need to strengthen the governance of antimicrobial stewardship in Ecuador through strategies that are contextually adapted, aligned with international recommendations, and grounded in a comprehensive multisectoral approach.

## Introduction

Antimicrobial resistance (AMR) constitutes a critical threat to global health [1], jeopardizing decades of progress in the treatment of infectious diseases. This challenge results in prolonged hospital stays, increased morbidity and mortality, and a substantial rise in healthcare costs[2–5]. Estimates suggest that AMR caused more than 1.27 million deaths in 2019, with the most severe impacts observed in low- and middle-income countries [6,7]. In Ecuador, several factors—such as socioeconomic inequality, limited and inefficient health infrastructure, self-medication, and the inappropriate use of antimicrobials in both human and animal health—exacerbate the situation [8–10]. These conditions promote selective pressure and facilitate the spread of resistant microorganisms [7]

The One Health (OH) approach recognizes the interdependence of human, animal, and environmental health, and promotes an integrated response to threats such as AMR, which transcend the boundaries of traditional health sectors [11]. In line with the OH approach and the *Global Action Plan on Antimicrobial Resistance* [12] a coordinated state strategy is required to address this critical public health threat. This include raising awareness and understanding through effective communication, education, and training; strengthening the evidence base via systematic surveillance and research; reducing infection incidence through improved sanitation, hygiene, a preventive measures; promoting the prudent use of antimicrobials in both human and animal health; and securing sustained investment in the development of new medicine, diagnostics, and vaccines. Reflecting these priorities, Ecuador implemented its *National Plan for the Prevention and Control of Antimicrobial Resistance (2019–2023)*. However, its implementation has been constrained by multiple structural barriers, including inadequate institutionalization of Ecuador’s National AMR Committee, insufficient budget allocation, weak intersectoral coordination, and limited political prioritization of the issue within the national agenda [13]. These challenges were further compounded by the impact of the COVID-19 pandemic, which diverted financial and operational resources and weakened AMR surveillance and response capacities [14]. Moreover, the shift in government from a *21^st^-century socialism* model to a neoliberal, market-oriented approach has reversed earlier gains, characterized by austerity measures, budged cuts, and institutional weakening. A recent political analysis notes that, since 2017, austerity policies have led to a severe reduction in public spending and the dismantling of social policies, public investment, and institutional capacity that had defined the previous decade [15]. These changes have had profound impacts on population health and the health system, including reduces access to essential services, deterioration in public health infrastructure, and worsened health outcomes, as evidenced during COVID-19 pandemic when austerity-driven cuts limited the health system’s capacity to respond effectively [16]. In Ecuador, the National AMR Committee was proposed in 2017 and legally formalized in 2020. It included representatives from the ministries of health, agriculture, environment, and higher education: however, implementation has been hindered by bureaucratic delays, scarce resources, political discontinuity, and weak multisectoral coordination.

In analyzing the AMR landscape in Ecuador from a OH perspective, this study assesses the governance and implementation of antimicrobial stewardship by examining key aspects such as the development of public policy, infrastructure, technical surveillance capacities and equity considerations. Structural and operational barriers, along with challenges in intersectoral coordination, are identified as major limitations to the implementation of the national plan. At the same time, the study highlights opportunities to strengthen mechanisms for collaboration, communication, and citizen engagement. Evaluating these factors is essential for the development of sustainable strategies that are contextually adapted to Ecuador and grounded in scientific evidence. Furthermore, this study aims to contribute to the local and regional dialogue on AMR in Latin America by emphasizing the urgency of more robust public policies that are equitable and intercultural, framed within a OH approach. Based on the analysis of the Ecuadorian case, a context-specific action framework is proposed to guide decision-making and inspire improvement processes in other countries in the region, thereby promoting the sustainability of interventions and their impact on public health.

## Materials and Methods

A qualitative approach was adopted to develop an environmental scan of the governance, infrastructure, public policies, and equity dimensions related to antimicrobial resistance (AMR) in Ecuador, under the OH approach. This methodology combined document analysis of public and private sector sources with semi-structured interviews conducted with key informants across various strategic sectors.

### Study Design

A case study of Ecuador was conducted within a broader comparative multi-country study involving Canada, Brazil, Rwanda, and Ecuador. Qualitative methods involved: (1) document analysis (environmental scan) and (2) semi-structured interviews. The objective was to identify barriers, gaps, and opportunities for improvement in strengthening the governance of AMR [17]

### Document Analysis

The document analysis was based on a systematic search strategy applied to institutional, academic, and legal sources in both Spanish and English. Search engines such as Google, Google Scholar and PubMED were used, along with the official websites of the Ministry of Public Health (MSP), Ministry of Agriculture and Livestock (MAG), Ministry of Environment, Water and Ecological Transition (MATE), National Agency for Health Regulation and Control (ARCSA), and various international organizations.

Keywords used included the terms “AMR”, “Antimicrobial resistance,” “antibiotic” or “antimicrobial,” “AMR Action Plan” or “National AMR Plan,” “Antimicrobial resistance in Ecuador” or “Antimicrobial use in Ecuador,” “One Health,” and its Spanish equivalents “Una Sola Salud” or “Una Salud.” Documents published between 2020 and 2024 were selected, provided they had direct relevance to the Ecuadorian context. Included materials comprised laws, regulations, executive decrees, technical reports, public policies, sectoral strategies, and both national and international scientific literature.

A total of 117 relevant documents were collected following a systematic screening process. Initially, 499 documents were identified related to global United Nations challenges, of which 240 addressed antimicrobial resistance (AMR) in general. From this subset, 137 documents specifically focused on AMR in Ecuador. After applying inclusion criteria, 117 documents were selected for in-depth analysis and uploaded into NVivo 13 software (version 1.7.2). The analysis was conducted through thematic coding, based on a structured codebook shared across the four participating countries. The main coding categories were: governance, infrastructure, public policy, and equity [18].

### Semi-structured interviews

A total of 23 semi-structured interviews were conducted with key informants selected through purposive sampling based on their direct experience in the design, implementation, or monitoring of AMR-related policies and plans. Participants represented human health, animal health, and environment sectors, academia, and civil society [19] (Table 1). The interview guide was developed and validated through expert review, and addressed the following thematic areas: governance structures, coordination mechanisms, implementation of the National AMR Plan, infrastructure gaps, and the application of equity-based approaches. Recruitment, and data collection began on March 1, 2024, and ended on June 15, 2024.

**Table 1.** Background of interview participants.

| Sector | Area of Expertise | Number of Participants | Percentage |
| --- | --- | --- | --- |
| University | AMR | 9 | 39.13 |
| Professional associations | AMR | 6 | 26.09 |
| Ministry of Health and affiliated agencies | AMR | 2 | 8.70 |
| Ministry of Agriculture and Livestock and agencies | AMR | 3 | 13.04 |
| Private institutions | AMR | 2 | 8.70 |
| Ministry of Enviroment | Climate change/ AMR | 1 | 4.35 |
| **Total** |  | **23** | **100.00** |

The interviews had an average duration of 45 minutes and were conducted in a hybrid format (in-person and virtual), depending on participants’ availability. Ethics approval was obtained from the Ethics Committee on Human Being Research, Universidad Central del Ecuador (CEISH Code “006-DOC-FMVZ-2023”). All participants were prospectively recruited and provided written informed consent prior to their participation. For in-person interviews, signed consent forms were obtained directly before the session. For participants interviewed virtually, the consent form was sent in advance by mail, signed and returned electronically before the interview took place. All sessions were audio-reordered with participants’ permission. No minors were included in this study.

Transcripts were analyzed using thematic coding in NVivo, with data triangulation performed through document analysis.

### Analytical Framework

Four analytical themes were defined as the interpretative framework for this study:

1. **Intersectoral governance analysis**: assessment of institutional mechanisms for coordination and multisectoral collaboration under the One Health approach.
2. **Situational analysis**: identification of objectives, positions, and levels of key stakeholder involvement.
3. **Transition toward One Health**: examination of facilitators and barriers for adoption of intersectoral strategies.
4. **Equity analysis using a GBA+ lens**: evaluation of how AMR-related policies incorporate gender equity, access to services, and the impact on vulnerable populations.

These themes were operationalized through nodes and sub-nodes coded in NVivo, structured within a thematic diagram that enabled linkage between documentary findings and qualitative interview data [20] (Fig 1).

**Figure 1.**
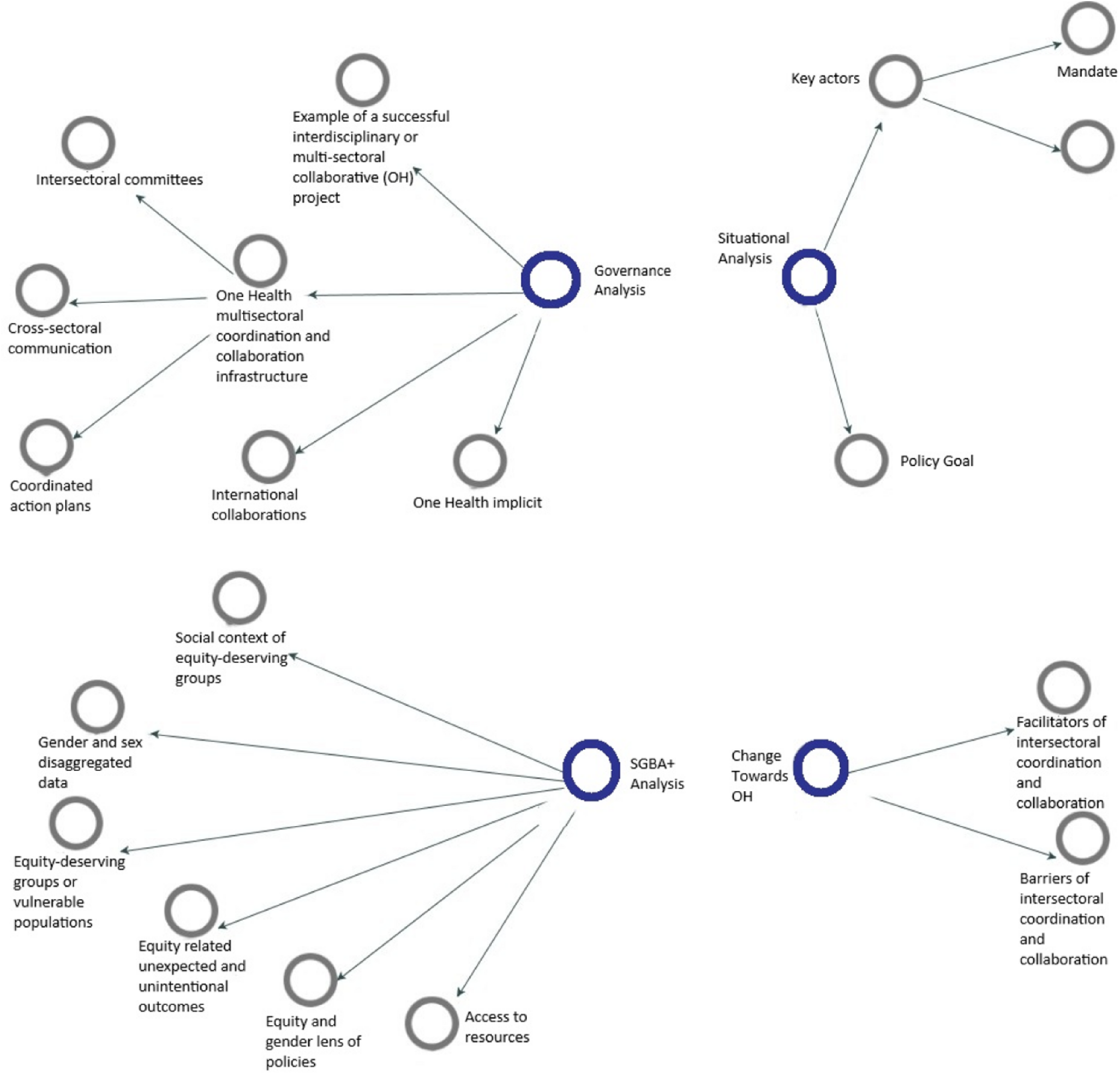
Codes used for the NVivo Analysis. **Note.** Diagram of the codes defined in NVivo software, showing thematic nodes (blue spheres) and sub-nodes (white spheres).

## Results

### Document analysis of AMR in Ecuador

The document analysis conducted using NVivo software enabled the coding of 117 documents from various sources, resulting in 742 coded files and 1,657 thematic references. The analyzed materials included institutional websites (n=42), government documents such as bulletins, gazettes, reports, manuals, and national plans (n=27), legal regulations (n=14), scientific articles (n=12), books (n=9), press releases (n=8), and academic theses (n=5). This information supported the construction of a hierarchical thematic map based on nodes aligned with the analytical framework: governance, infrastructure, public policy, and equity. The distribution of codes and sources by node is presented in Table 2. Further details are available in Annex 1.

**Table 2.** Results of thematic coding of AMR-related documents.

| Main node | Files* | References** | Documents Types (summary) |
| --- | --- | --- | --- |
| Governance analysis | 100 | 288 | Government documents, legal regulations, institutional websites |
| Situational analysis | 90 | 293 | Regulations, policy documents, academic articles, theses |
| Transition toward One Health | 33 | 71 | Intersectoral reports, scientific articles, theses |
| Gender-Based analysis (GBA+) | 24 | 77 | Regulations, scientific literature, documents with an equity approach |

\* Files: Total number of documents coded under one or more nodes.
\*\* References: Number of document excerpts coded.

### Governance and intersectoral Collaboration in One Health

Findings from the interviews with 23 key stakeholders revealed contrasting perceptions regarding OH governance. While most (96 percent) participants reported being familiar with the term “One Health,” only half (48 percent) identified the existence of concrete governance infrastructure enabling this approach. Moreover, just thirty percent of key informants (KIs) were aware of intersectoral communication tools implemented within their institutional practice. Multisectoral collaboration on AMR was perceived as limited: seventy-four percent indicated that they lacked a clear network of inter-institutional allies or contacts. Weak articulation of the One Health approach among responsible entities was found to hinder the effective implementation of the *National Plan for the Prevention and Control of Antimicrobial Resistance (2019–2023)* [13], as evidenced by both the documents analyzed and the perceptions gathered through interviews.

The lack of formal coordination mechanisms between ministries and agencies limits the effectiveness of the national response. Although a national AMR committee was officially established to support the implementation of the National Plan, it is currently inactive. Such fragmentation—confirmed by the majority of interview participants—is further exacerbated by the lack of effective regulation and oversight regarding the rational use of antimicrobials across all sectors.

### Equity and Gender in Health approaches

Regarding equity, participants identified traditionally excluded groups as priority populations, including rural communities, low-income populations, migrants, small-scale producers, and immunocompromised individuals. When probed, some respondents explicitly referred to gender equity, others to broader social and economic equity, and a few to both dimensions. However, less than five percent of interviewees believed that either gender equity or general equity is effectively integrated into current AMR policies. Most respondents (74 percent) indicated that these considerations, regardless of type, are either absent or only marginally addressed, while the remaining participants expressed a lack of knowledge on the matter, suggesting that responses may partially reflect limited awareness among respondents. These findings highlight the need to systematically integrated both gender equity and broader equity considerations into regulatory and institutional frameworks.

### Barriers and facilitators

The interviews also identified multiple barriers to effective collaboration under the One Health approach. The most frequently mentioned obstacles included bureaucracy, lack of political will, limited technical understanding of the approach, scarcity of financial resources, corruption, and resistance to change.

One of the main barriers identified is the fragility of the infrastructure for epidemiological surveillance. The Integrated Epidemiological Surveillance System (SIVE) [21,22] requires technological modernization and the establishment of interoperable mechanisms across sectors. Moreover, reference laboratories face persistent challenges, including insufficient funding, a shortage of trained personnel, and inadequate equipment—factors that undermine the generation of reliable data for decision-making. Without a robust and coordinated surveillance system, the development of effective AMR policies is severely compromised.

Community misinformation and the irrational use of antimicrobials remain persistent challenges, particularly among vulnerable populations and in rural areas. Self-medication and unrestricted access to antibiotics—widely documented in the literature [23,24] —are further exacerbated by sporadic and poorly coordinated awareness campaigns. The findings of this study underscore the urgent need for sustained communication strategies, coordinated with academic institutions and community-based organizations, to promote the rational use of antimicrobials.

Conversely, the main facilitating factors cited were improved intersectoral communication, the organization of meetings and workshops, technical training, political leadership, and the inclusion of civil society actors. Over ninety percent agreed that promoting stronger intersectoral collaboration is both feasible and desirable within the country. Table 3 summarizes the most relevant responses provided by interview participants. Detailed findings are presented in Annex 2.

**Table 3.** Key informant interview results on governance, equity and intersectoral barriers.

| Category | Key response | Percentage (%) |
| --- | --- | --- |
| Familiarity with One Health | Yes | 95.65 |
| Use of intersectoral communication tools | Yes | 30.43 |
| Presence of an AMR contact network | No | 73.91 |
| Feasibility of multisectoral collaboration | Yes | 91.30 |
| Equity integrated into AMR policies | Yes | 4.35 |
| Most frequently cited barrier | Political will | 21.74 |
| Most frequently cited facilitator | Meeting/workshop/political engagement | 21.74 |

Table 4 presents the main structural barriers and facilitating factors identified through documents analysis and key informant interviews regarding the implementation of the OH approach to address AMR in Ecuador. Six key barriers emerged including: limited intersectoral coordination, insufficient budged allocations, poor infrastructure, high staff turnover, lack of political prioritization, and limited public education coupled with high levels of self-medications. In contrast, six facilitating factors were identified, such as: the existence of a national AMR surveillance system, a favorable legal framework, willingness to collaborate among stakeholders, adoption of international programs, the active role of academia in policy development, and the adaptability of international guidelines and protocols to the national context.

**Table 4.** Barriers and facilitators for addressing AMR in Ecuador.

| Barriers | Facilitators |
| --- | --- |
| Lack of intersectoral integration and coordination: absence of joint working groups across human, animal, and environmental health. | AMR surveillance system: the CRN-RAM of INSPI provides relevant data that fosters collaboration. |
| Insufficient budget due to fiscal crisis, drug trafficking, and public debt. | Favorable legal framework: decrees and resolutions support plans under the One Health approach. |
| Poor infrastructure: underperforming laboratories, budget cuts, and insufficient personnel. | Willingness to collaborate: identified among public and academic sector stakeholders. |
| High staff turnover and lack of ongoing training programs. | Adoption of international programs such as Antimicrobial Stewardship Programs (PROA). |
| Lack of political will: AMR is not prioritized on government agendas. | Role of academia: scientific evidence supports public policy development. |
| Limited public education and high levels of self-medication. | International guidelines and protocols can be adapted to the national context. |

## Discussion

Relevance of the One Health approach as a strategic framework for addressing antimicrobial resistance (AMR) are reaffirmed by the study findings. This integrative perspective enables a coherent understanding of the interconnections between human, animal, plant, and environmental health—critical elements in AMR control. However, Ecuador faces significant limitations in the implementation of this approach, particularly in terms of limited political will, fragmented governance and interinstitutional coordination.

The analysis also reveals the insufficient incorporation of an equity-based approach— particularly with regard to gender—within public policies related to AMR. Groups such as rural women, immunocompromised individuals, and people living in poverty face significant barriers in accessing appropriate diagnostics and treatment. Nevertheless, this finding highlights the urgent need to mainstream the GBA+ (Gender-Based Analysis Plus) approach into health regulations and programs.

To enhance the national response, the establishment of a structured governance framework that integrates human, animal, and environmental health—while also engaging academic institutions and civil society—is essential. This would involve strengthening the AMR Committee with adequate resources, a strong mandate, and the active participation of academia and civil society organizations. A national AMR coordination body could centralize efforts, optimize resources, and ensure policy continuity. Moreover, the adaptation of international models—such as Antimicrobial Stewardship Programs (PROA)—could contribute to the technical capacity-building of national institutions by tailoring protocols, training modules, and monitoring tools to the local epidemiological context and resource availability *[25,26]*.

The Table 4 identifies and summarizes our findings concerning six key structural barriers and six facilitating factors that present concrete opportunities to improve the articulation of the OH approach in addressing AMR in Ecuador. These findings also reveal important limitations that should be acknowledged, including inadequate infrastructure, insufficient budget allocations, high staff turnover, lack of ongoing training programs, limited public education, high levels of self-medications, and the absence of AMR as a governmental priority. Recognizing these constrains is essential to contextualize the feasibility and scope of proposed interventions.

In the Latin American context, countries face similar structural barriers to AMR control, such as limited laboratory capacity, fragmented governance, and underfunded public health systems. However, several regional experiences demonstrate that these obstacles can be overcome. In Argentina, the implementation of antimicrobial stewardship programs (ASPs) in hospitals -supported by national guidelines, preauthorization protocols, and regular audits- has reduced inappropriate antimicrobial use by up to 20% and lowered infections-related mortality rates [27]. Brazil and Colombia have strengthened AMR governance by establishing multisectoral coordination committees with clear mandates, designated budget lines, and integration of the OH approach [28]. Regional initiatives such as the Latin American and Caribbean Network for Antimicrobial Resistance Surveillance (ReLAVRA+), coordinated by PAHO, have enhanced data standardization, strengthened laboratory capacity, and fostered cross-border collaborations [29]. Positioning Ecuador’s efforts within this regional landscape may promote an enabling environment for transferable strategies, such as scaling up hospital-levels ASPs adapted to local epidemiology context, formalizing and resourcing the National ARM Committee, leveraging regional networks for technical support, and implementing target public education campaigns to address self-medication. These experiences underscore the feasibility of overcoming structural barriers through evidence-based, context-specific interventions.

## Conclusions

Ecuador faces structural, institutional, and social challenges that constrain its national response to antimicrobial resistance (AMR). This study revealed fragmented governance, weaknesses in epidemiological surveillance systems, and insufficient intersectoral coordination. Although a *National Plan for the Prevention and Control of Antimicrobial Resistance (2019–2023)* was developed, limited institutionalization, the absence of effective coordination mechanisms, and weak regulatory compliance have hindered its implementation—undermining Ecuador’s capacity to respond to this global health threat.

This study documented a deterioration of governance capacities, interoperability of information systems, laboratory infrastructure, and budget allocations for AMR surveillance programs in Ecuador, compared to the conditions established under the prior centrist administration that developed responsive public sector institutions. The analysis also revealed a reduction in community education efforts on the rational use of antimicrobials, which may be contributing to persistent self-medication practices, particularly among vulnerable and rural populations.

Overcoming these barriers requires sustained political commitment, supported by a national governance structure that coordinates efforts across government, academia, professional and private sector associations, civil society and AMR Committees. The creation or strengthening of a regulatory body to centralize AMR initiatives would improve intersectoral coordination, optimize resource allocation, and ensure the continuity of prevention and control strategies.

Finally, integrating the One Health approach as a cross-cutting axis in policy formulation will enable Ecuador to advance toward a more efficient, sustainable, and inclusive national response to AMR. This transformation is critical not only for protecting public health, but also for safeguarding food security, environmental sustainability, and the resilience of the health system in the face of future threats.

## Data Availability

All relevant data supporting the findings of this study are fully available within the manuscript. Aditional information is available from the authors upon reasonable request.

## Acknowledgments

We thanks all expert participants who voluntarily provided valuable information to understand AMR situation in Ecuador. We also acknowledge the financial support of the “New Frontiers in Research Fund. Especial Call 2022 (code NFRFR-2020-00492) – Canadá, administered by York University and the “Empresa Publica de la Universidad Central del Ecuador (EP-UCE)”

## Author contributions

**Conceptualization:** Mary Wiktorowicz, Arne Ruckert, Raphael Aguiar, Phaedra Henley, Mayumi Duarte Wakimoto, Richar Rodriguez-Hidalgo.

**Investigation**: Richar Rodriguez-Hidalgo, Linette Jácome, Diana Paz.

**Methodology**: Linette Jácome, Diana Paz, Richar Rodriguez-Hidalgo, Arne Ruckert, Raphael Aguiar.

**Project administration**: Richar Rodriguez-Hidalgo, Mary Wiktorowicz.

**Resources**: Linette Jácome, Diana Paz.

**Supervision**: Richar Rodriguez-Hidalgo.

**Validation**: Mary Wiktorowicz, Arne Ruckert, Raphael Aguiar, Phaedra Henley, Mayumi Duarte Wakimoto.

**Visualization**: Richar Rodriguez-Hidalgo.

**Writing-original draft**: Richar Rodriguez-Hidalgo, Linette Jácome.

**Writing-review & editing**: Mary Wiktorowicz, Arne Ruckert, Raphael Aguiar, Phaedra Henley, Mayumi Duarte Wakimoto.

## Annexes or Supporting Information

## Annex 1

### Results of Thematic Coding of AMR-Related Documents

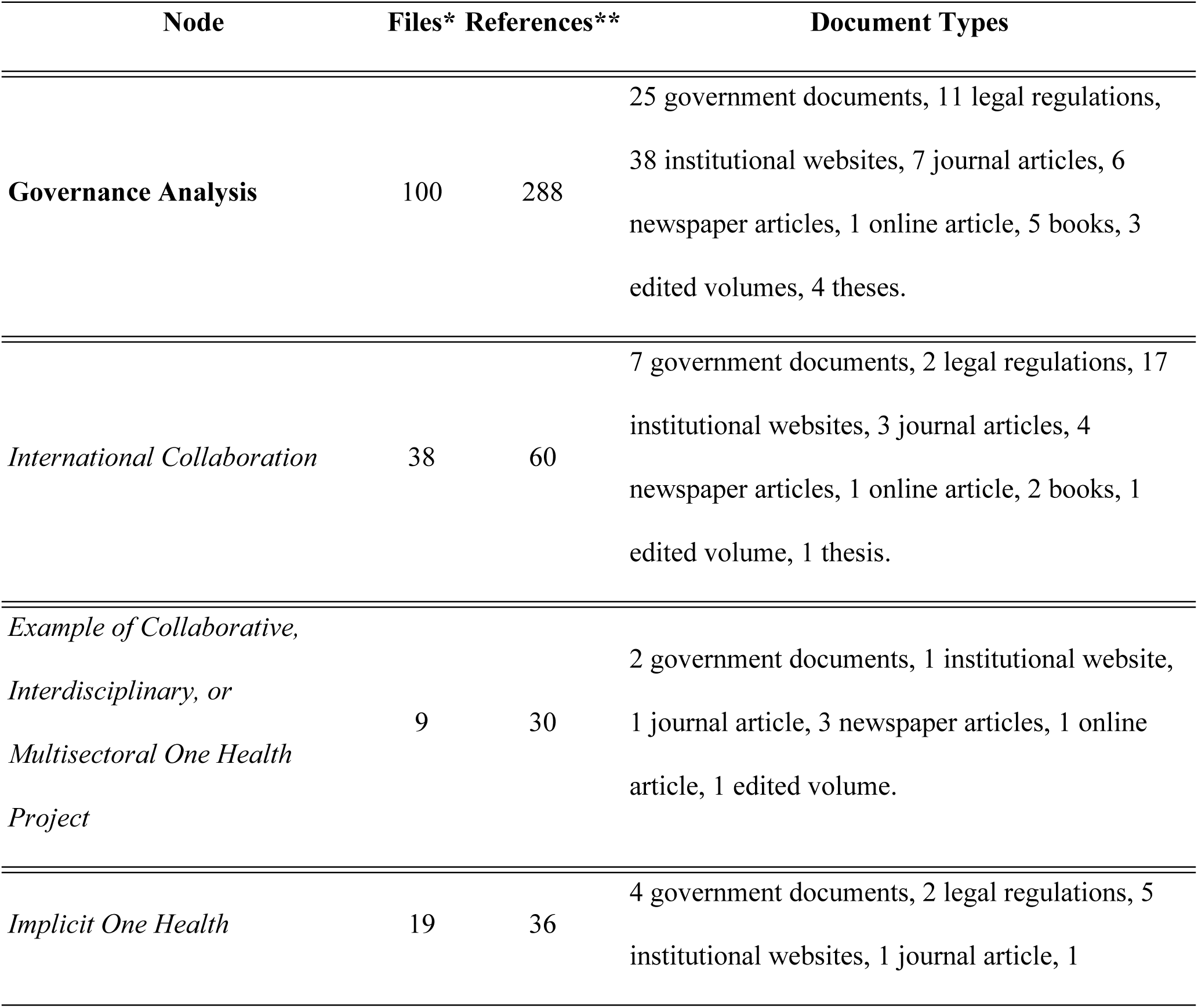
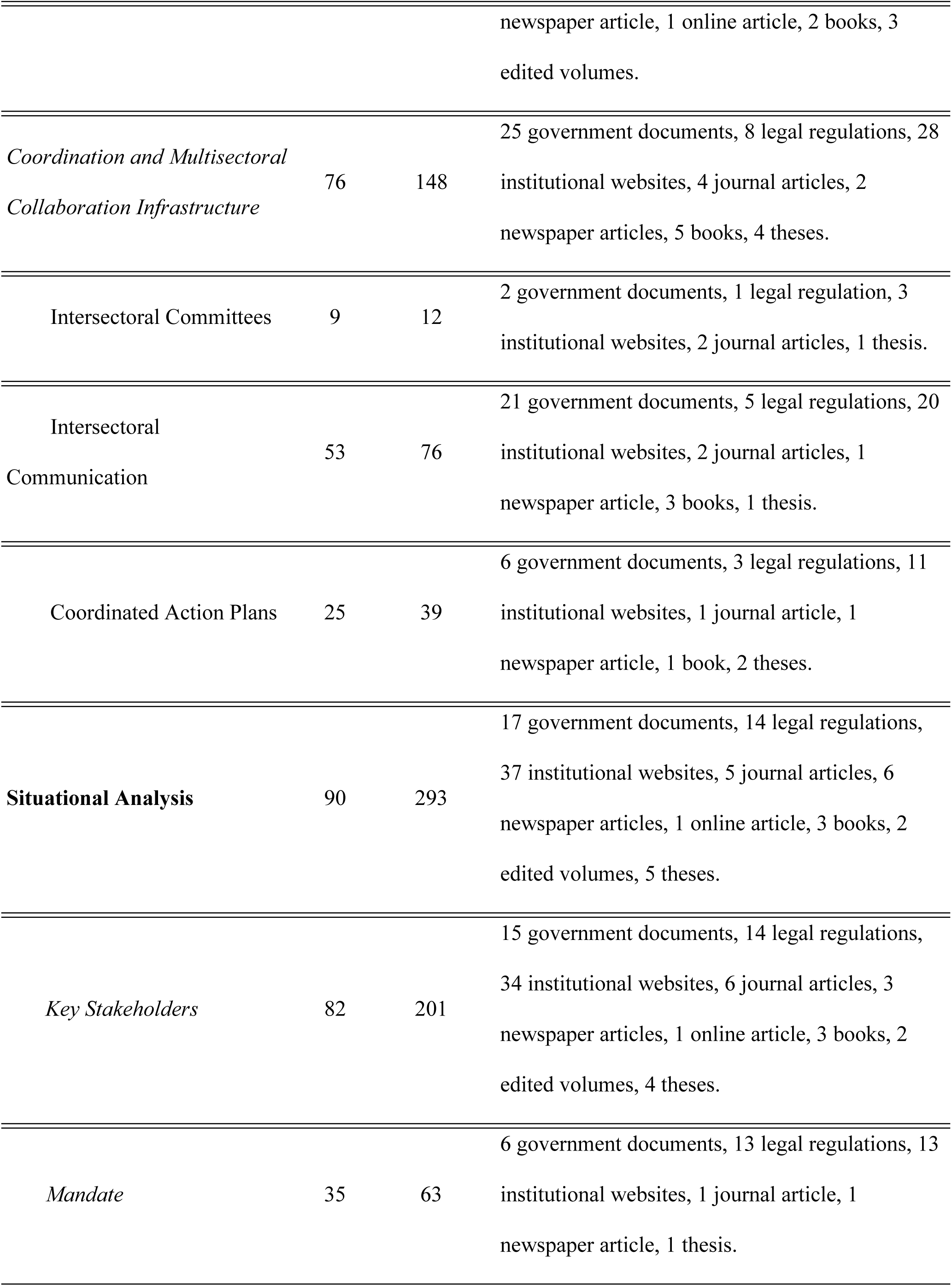
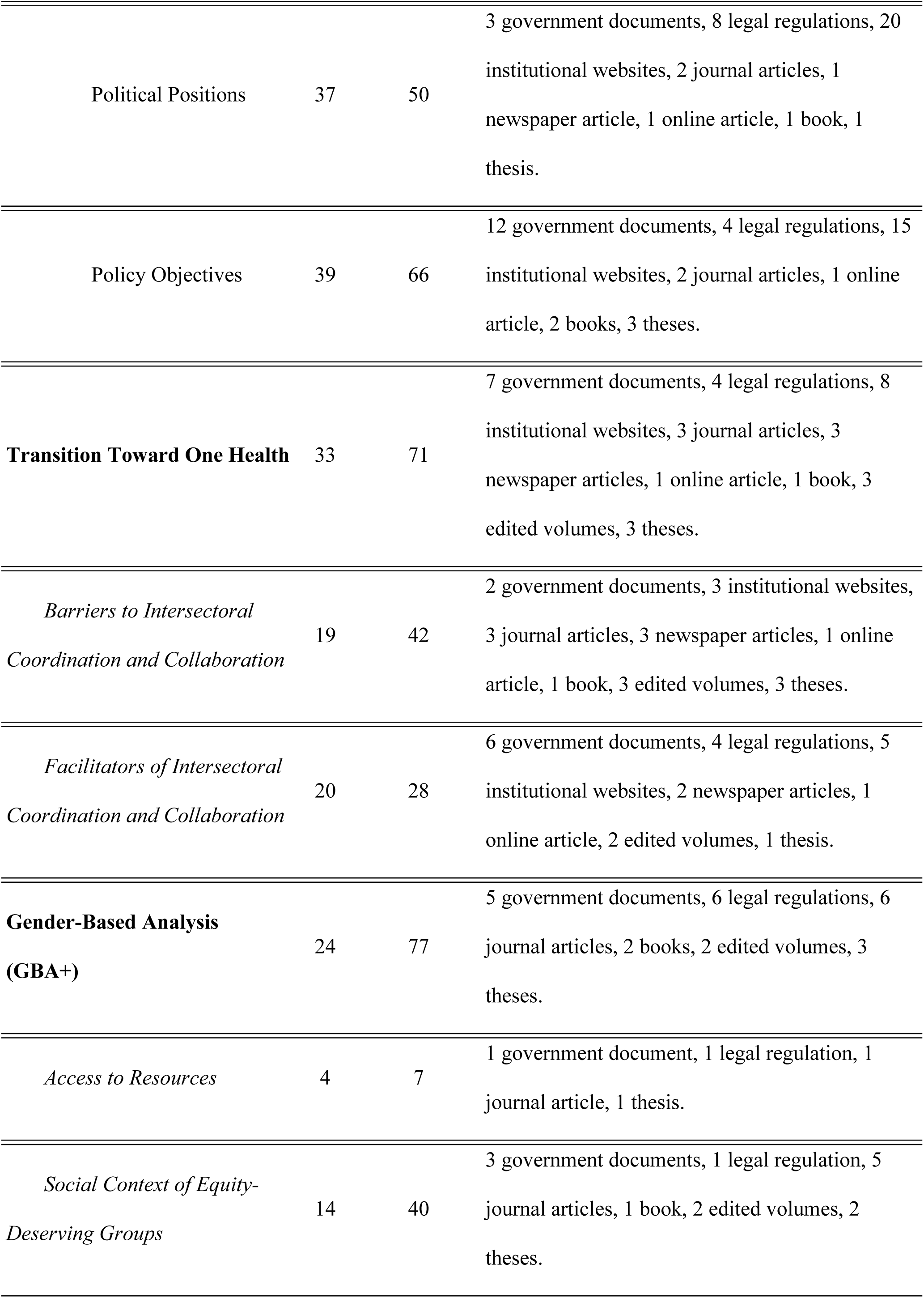
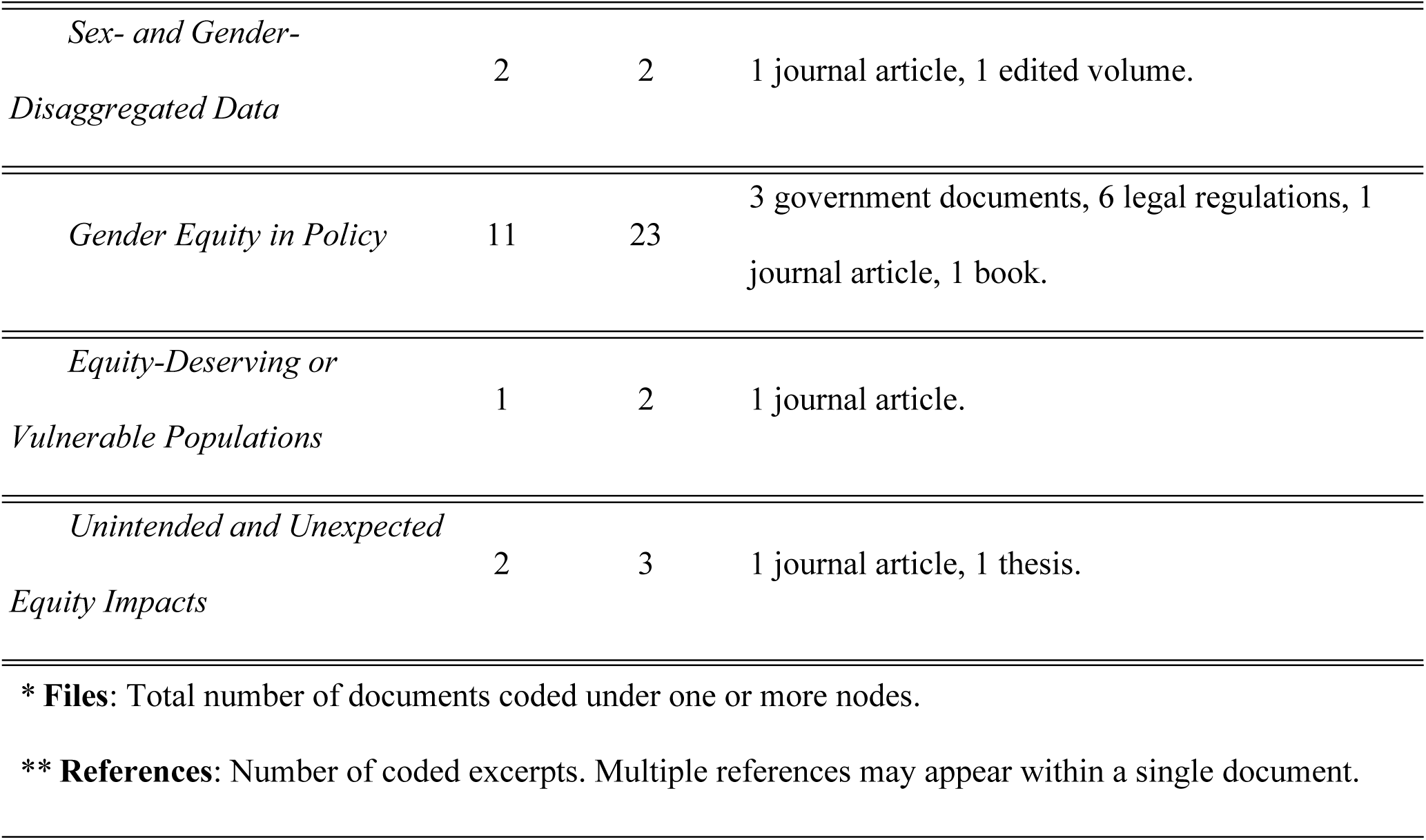

| Node | Files* | References** | Document Types |
| --- | --- | --- | --- |
| **Governance Analysis** | 100 | 288 | 25 government documents, 11 legal regulations, 38 institutional websites, 7 journal articles, 6 newspaper articles, 1 online article, 5 books, 3 edited volumes, 4 theses. |
| *International Collaboration* | 38 | 60 | 7 government documents, 2 legal regulations, 17 institutional websites, 3 journal articles, 4 newspaper articles, 1 online article, 2 books, 1 edited volume, 1 thesis. |
| *Example of Collaborative, Interdisciplinary, or Multisectoral One Health Project* | 9 | 30 | 2 government documents, 1 institutional website, 1 journal article, 3 newspaper articles, 1 online article, 1 edited volume. |
| *Implicit One Health* | 19 | 36 | 4 government documents, 2 legal regulations, 5 institutional websites, 1 journal article, 1 |

| **Node** | **Files\*** | **References\*\*** | **Document Types** |
| --- | --- | --- | --- |
|  |  |  | newspaper article, 1 online article, 2 books, 3 edited volumes. |
| *Coordination and Multisectoral Collaboration Infrastructure* | 76 | 148 | 25 government documents, 8 legal regulations, 28 institutional websites, 4 journal articles, 2 newspaper articles, 5 books, 4 theses. |
| Intersectoral Committees | 9 | 12 | 2 government documents, 1 legal regulation, 3 institutional websites, 2 journal articles, 1 thesis. |
| Intersectoral Communication | 53 | 76 | 21 government documents, 5 legal regulations, 20 institutional websites, 2 journal articles, 1 newspaper article, 3 books, 1 thesis. |
| Coordinated Action Plans | 25 | 39 | 6 government documents, 3 legal regulations, 11 institutional websites, 1 journal article, 1 newspaper article, 1 book, 2 theses. |
| **Situational Analysis** | 90 | 293 | 17 government documents, 14 legal regulations, 37 institutional websites, 5 journal articles, 6 newspaper articles, 1 online article, 3 books, 2 edited volumes, 5 theses. |
| *Key Stakeholders* | 82 | 201 | 15 government documents, 14 legal regulations, 34 institutional websites, 6 journal articles, 3 newspaper articles, 1 online article, 3 books, 2 edited volumes, 4 theses. |
| *Mandate* | 35 | 63 | 6 government documents, 13 legal regulations, 13 institutional websites, 1 journal article, 1 newspaper article, 1 thesis. |
| Political Positions | 37 | 50 | 3 government documents, 8 legal regulations, 20 institutional websites, 2 journal articles, 1 newspaper article, 1 online article, 1 book, 1 thesis. |
| Policy Objectives | 39 | 66 | 12 government documents, 4 legal regulations, 15 institutional websites, 2 journal articles, 1 online article, 2 books, 3 theses. |
| **Transition Toward One Health** | 33 | 71 | 7 government documents, 4 legal regulations, 8 institutional websites, 3 journal articles, 3 newspaper articles, 1 online article, 1 book, 3 edited volumes, 3 theses. |
| *Barriers to Intersectoral Coordination and Collaboration* | 19 | 42 | 2 government documents, 3 institutional websites, 3 journal articles, 3 newspaper articles, 1 online article, 1 book, 3 edited volumes, 3 theses. |
| *Facilitators of Intersectoral Coordination and Collaboration* | 20 | 28 | 6 government documents, 4 legal regulations, 5 institutional websites, 2 newspaper articles, 1 online article, 2 edited volumes, 1 thesis. |
| **Gender-Based Analysis (GBA+)** | 24 | 77 | 5 government documents, 6 legal regulations, 6 journal articles, 2 books, 2 edited volumes, 3 theses. |
| *Access to Resources* | 4 | 7 | 1 government document, 1 legal regulation, 1 journal article, 1 thesis. |
| *Social Context of Equity-Deserving Groups* | 14 | 40 | 3 government documents, 1 legal regulation, 5 journal articles, 1 book, 2 edited volumes, 2 theses. |
| *Sex- and Gender-Disaggregated Data* | 2 | 2 | 1 journal article, 1 edited volume. |
| *Gender Equity in Policy* | 11 | 23 | 3 government documents, 6 legal regulations, 1 journal article, 1 book. |
| *Equity-Deserving or Vulnerable Populations* | 1 | 2 | 1 journal article. |
| *Unintended and Unexpected Equity Impacts* | 2 | 3 | 1 journal article, 1 thesis. |

\* **Files:** Total number of documents coded under one or more nodes.
\*\* **References:** Number of coded excerpts. Multiple references may appear within a single document.

## Annex 2

### Results of Key Informant Interviews on Governance, Equity, and Intersectoral Barriers

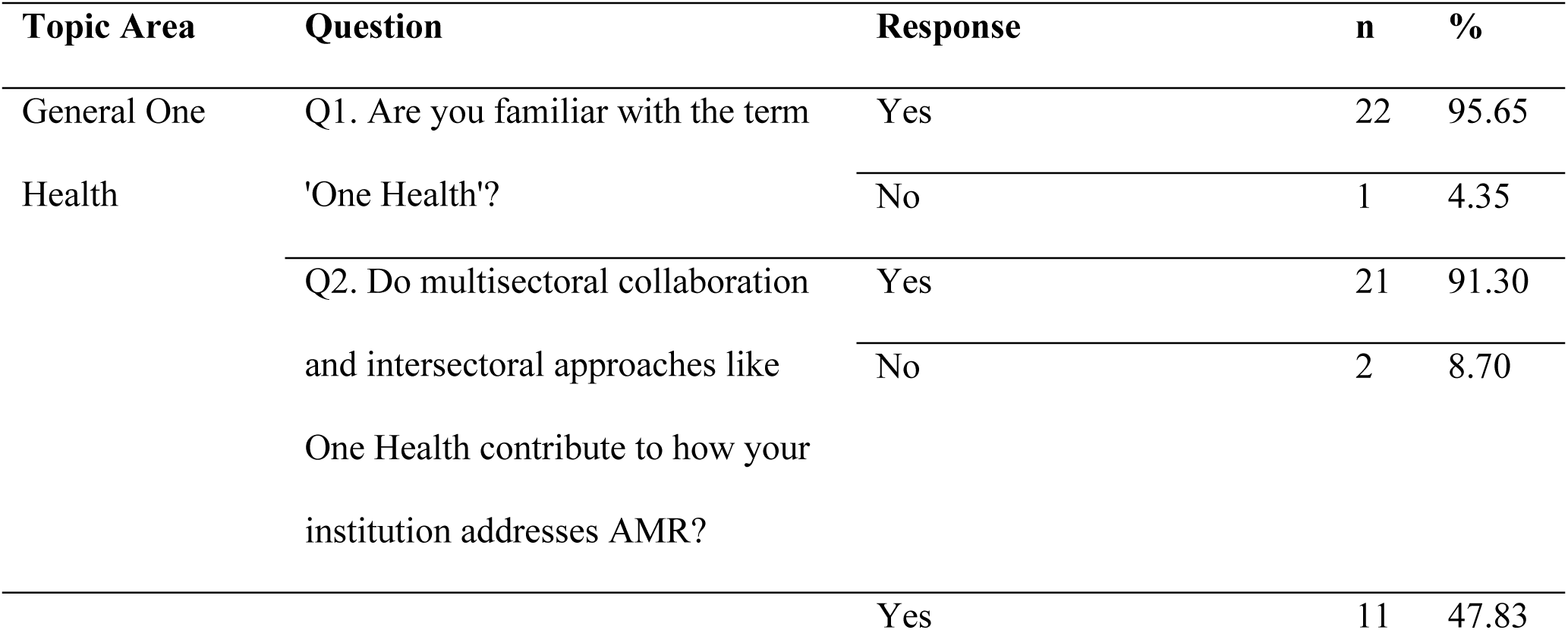
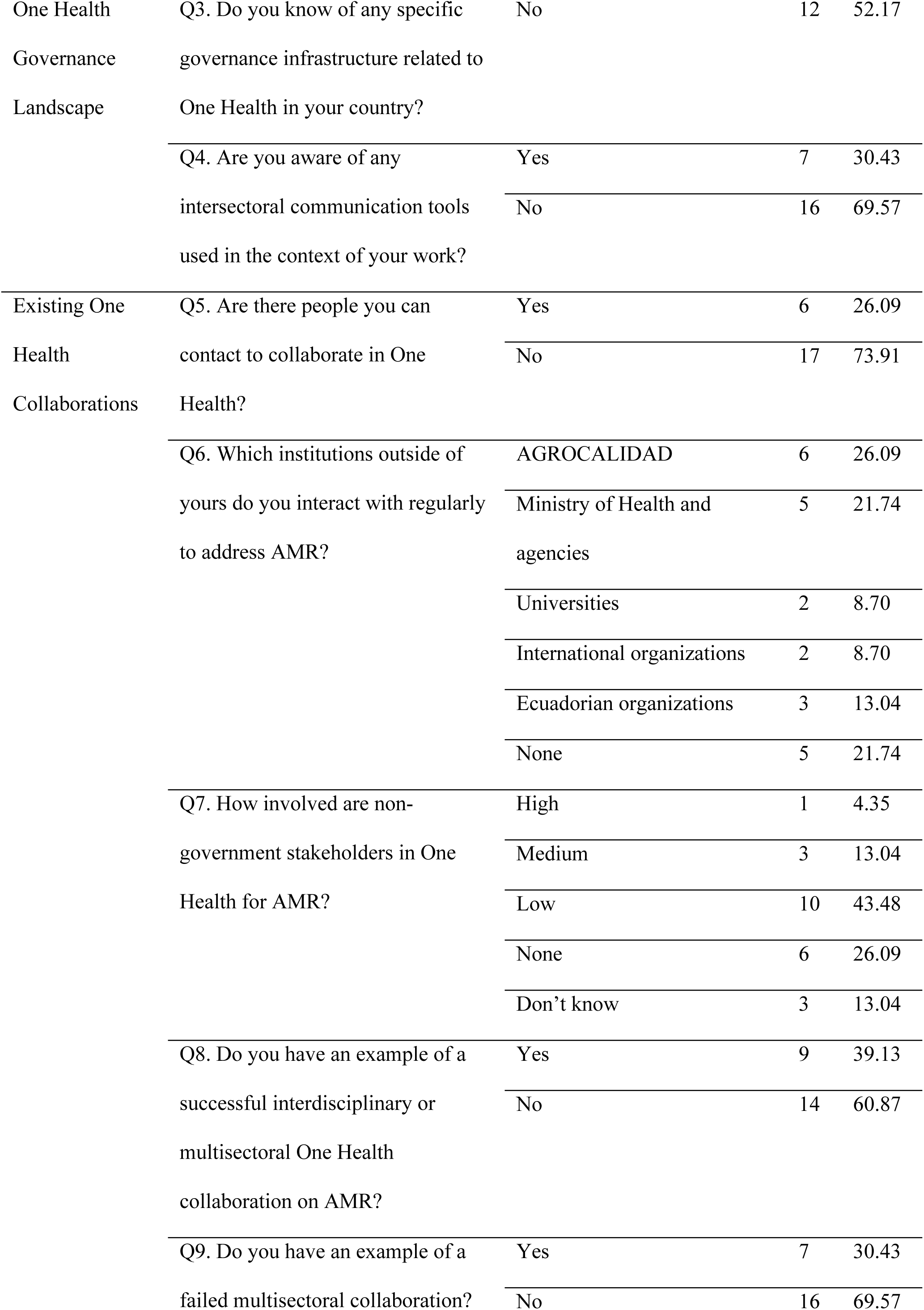
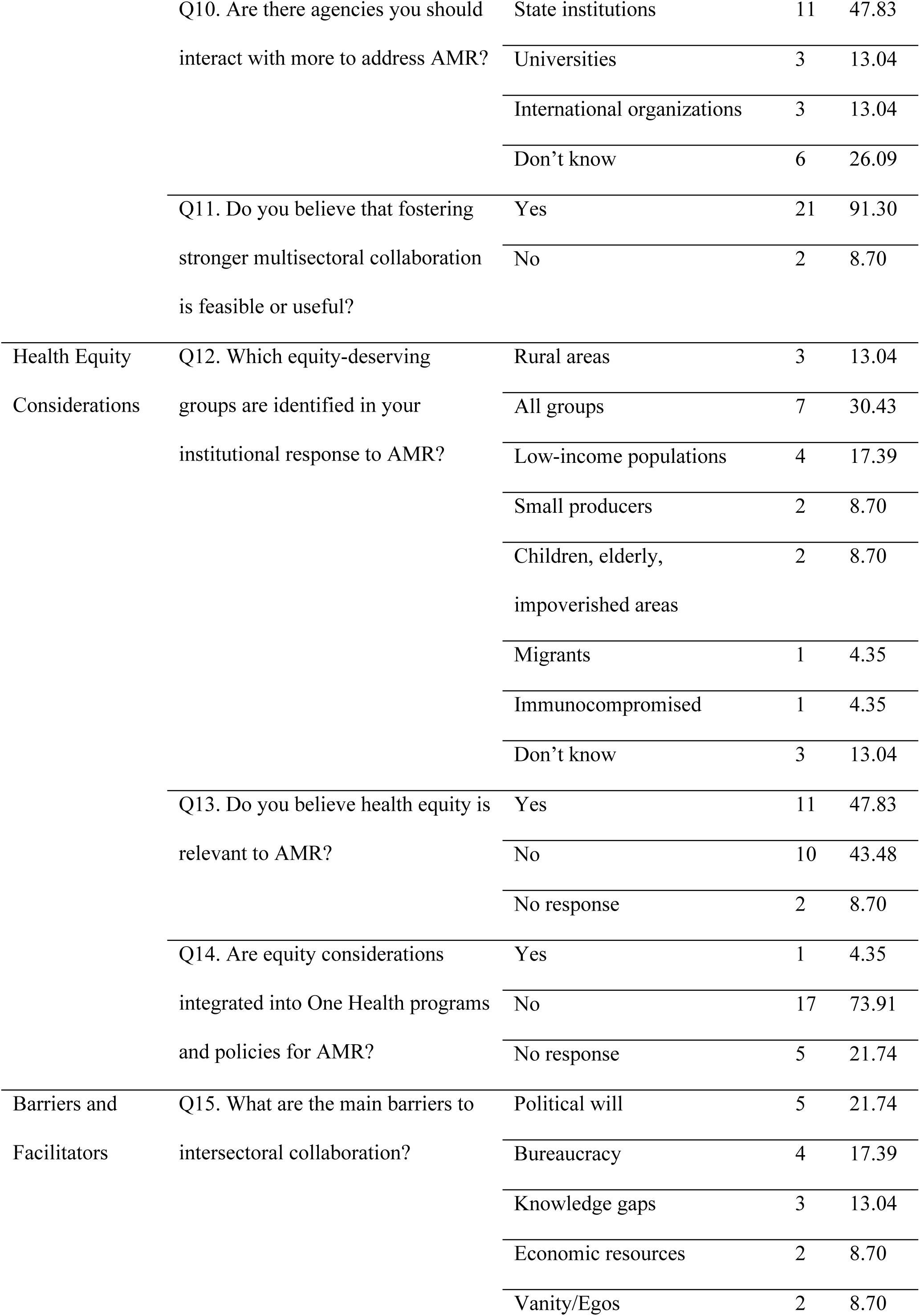
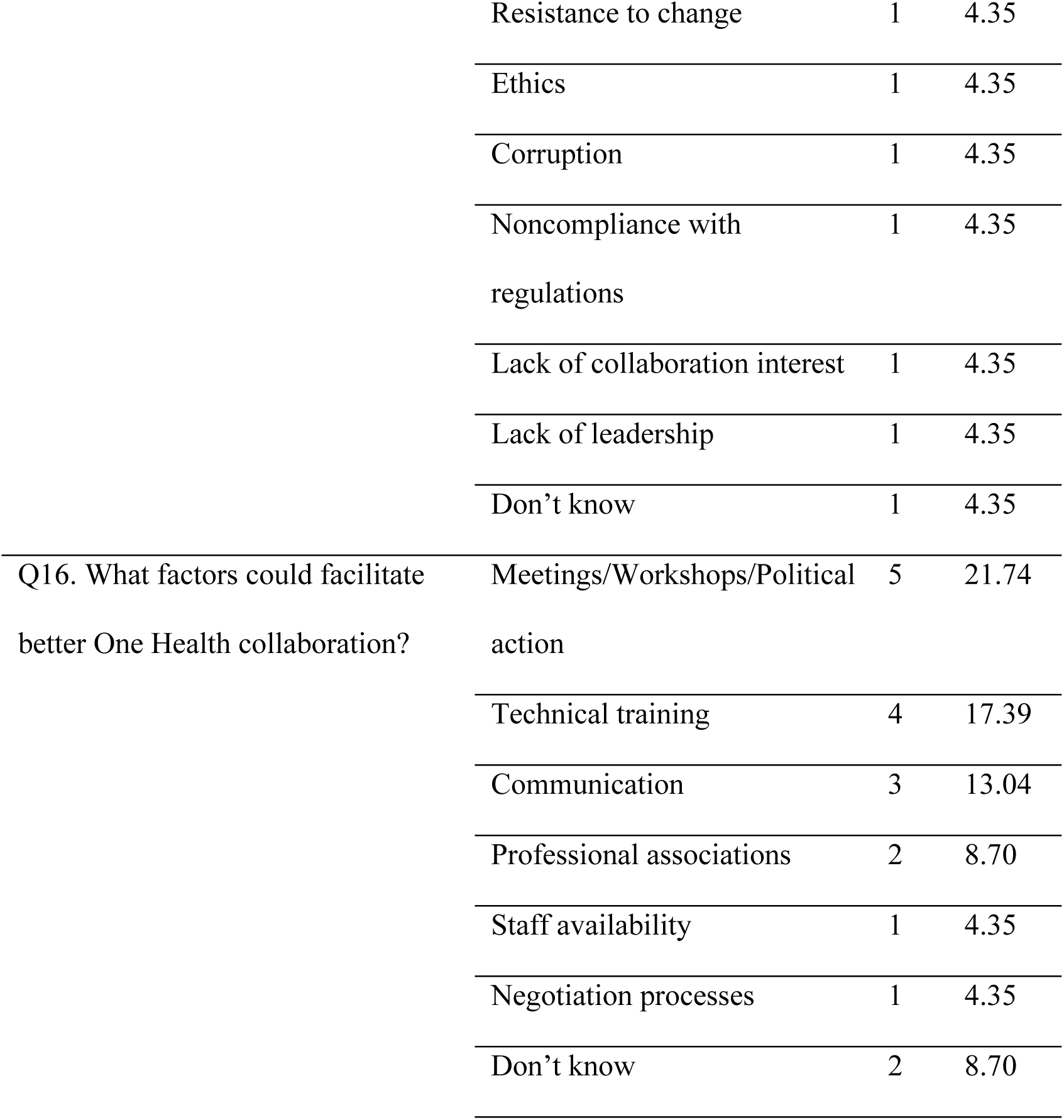

## Notes

### Competing Interest Statement

The authors have declared no competing interest.

### Funding Statement

This research was funded by the New Frontiers in Research Fund - Special Call 2022 (Application ID: NFRFR-2022-00492), administrated by York University (Canada). The project is titled "Governance of One Health challenges: Fostering collaboration." The funder had no role in this study design, data collection and analysis, decision to publish, or preparation of manuscript. Principal and co-applicants include researchers from York University (Canada), University of Global Health Equity (Rwanda), University of Ottawa (Canada), Oswaldo Cruz Foundation - Evadro Chagas National Institute of Infectious Diseases (Brazil), and Universidad Central del Ecuador - Instituto de Investigaciones en Zoonosis - CIZ.

### Author Declarations

The study protocol was reviewed and approved by the Ethics Committee for Research on Human Beings of the Universidad Central del Ecuador (Approval No. 006-DOC-FMVZ-2023, July 25, 2023)

## References

1. OMS. Diez cuestiones de salud que la OMS abordará este año. In: Organización Mundial de la Salud [Internet]. 2019 [cited 3 Feb 2025]. Available: https://www.who.int/es/news-room/spotlight/ten-threats-to-global-health-in-2019

2. OMS. Resistencia a los antimicrobianos. In: Organización Mundial de la Salud [Internet]. 2021 [cited 7 May 2024]. Available: https://www.who.int/es/news-room/fact-sheets/detail/antimicrobial-resistance

3. OMS. La resistencia a los antimicrobianos y el Marco de Cooperación de las Naciones Unidas para el Desarrollo Sostenible. Organización Mundial de Sanidad Animal, editor. Organización Mundial de la Salud. Organización Mundial de Sanidad Animal; 2021. Available: https://openknowledge.fao.org/server/api/core/bitstreams/5080b8aa-91b9-4087-be4a-3c2eb5d24df0/content

4. O’Neill. Tackling drug-resistant infections globally: final report and recommendations the review on antimicrobial resistance. 2016. Available: https://amr-review.org/sites/default/files/160518_Final%20paper_with%20cover.pdf

5. CDC. Antibiotic resistance threats in the United States, 2019. Atlanta, Georgia; 2019 Nov. doi:10.15620/cdc:82532

6. ReAct Latinoamérica. Comunidades Empoderadas frente la RAM: Nuestro Planeta, Nuestra Salud. In: ReAct Latinoamérica [Internet]. 2024. Available: https://reactlat.org/comunidad-antibioticos/comunidades-empoderadas-frente-la-ram-nuestro-planeta-nuestra-salud/

7. Murray CJ, Ikuta KS, Sharara F, Swetschinski L, Robles Aguilar G, Gray A, et al. Global burden of bacterial antimicrobial resistance in 2019: a systematic analysis. The Lancet. 2022;399: 629–655. doi:10.1016/S0140-6736(21)02724-0

8. Espinosa V, Acuña Cecilia, De la Torre D, Tambini G. La reforma en salud del Ecuador. Rev Panamericana de Salud Pública. 2017;41: 449–461. Available: http://iris.paho.org/xmlui/bitstream/handle/123456789/34061/v41a962017.pdf

9. Goyes-Baca MJ, Sacon-Espinoza MR, Poveda-Paredes FX. Manejo del sistema de salud de Ecuador frente a la resistencia antimicrobiana Management. Revista Información Científica. 2023;102: 1–15. doi:10.5281/zenodo.7545370

10. MSP. Plan Nacional para la prevención y control de la resistencia antimicrobiana. 2019 [cited 29 May 2025]. Available: https://www.salud.gob.ec/wp-content/uploads/2019/10/Plan-Nacional-para-la-prevenci%C3%B3n-y-control-de-la-resistencia-antimicrobiana_2019_compressed.pdf

11. de Kraker MEA, Stewardson AJ, Harbarth S. Will 10 Million People Die a Year due to Antimicrobial Resistance by 2050? PLoS Med. 2016;13. doi:10.1371/JOURNAL.PMED.1002184,

12. OMS. Plan De Acción Mundial Sobre La Resistencia a Los Antimicrobianos. Organización Mundial de la Salud. 2016. Available: https://www.who.int/es/publications/i/item/9789241509763

13. MSP. Plan Nacional para la prevención y control de la resistencia antimicrobiana. Ministerio de Salud Pública del Ecuador. 2019; 1–33. Available: https://www.salud.gob.ec/wp-content/uploads/2019/10/Plan-Nacional-para-la-prevención-y-control-de-la-resistencia-antimicrobiana_2019_compressed.pdf

14. OPS. El impacto de la COVID-19 en la resistencia antimicrobiana. In: Organización Panamericana de la Salud [Internet]. 2021 [cited 19 Mar 2025]. Available: https://www.paho.org/es/noticias/17-11-2021-impacto-covid-19-resistencia-antimicrobiana

15. Johnston J, Vasic-Lalovic I. Ecuador: A Decade of Progress, Undone. Washington DC; 2023.

16. Ortiz I, Cummins M. Austerity: The New Normal - A Renewed Washington Consensus 2010-24. SSRN Electronic Journal. 2019. doi:10.2139/ssrn.3523562

17. Lungu M. The Coding Manual for Qualitative Researchers. American Journal of Qualitative Research. 2022;6: 232–237. doi:10.29333/AJQR/12085

18. Braun V, Clarke V. Using thematic analysis in psychology. Qual Res Psychol. 2006;3: 77–101. doi:10.1191/1478088706QP063OA

19. Krueger RA., Casey MAnne. Focus groups : a practical guide for applied research. 2015 [cited 29 May 2025]. Available: https://books.google.com/books/about/Focus_Groups.html?id=APtDBAAAQBAJ

20. Glaser BG, Strauss AL. The Discovery of Grounded Theory. Routledge; 2017. doi:10.4324/9780203793206

21. MSP. Normas del sistema integrado de vigilancia epidemiológica del ecuador (SIVE). Ministerio de Salud Pública del Ecuador. 2012; 1–25. Available: https://aplicaciones.msp.gob.ec/salud/archivosdigitales/documentosDirecciones/dnn/archivos/NORMAsive8-04-2013.pdf

22. MSP. Sistema Integrado de Vigilancia Epidemiológica Norma técnica Sistema Integrado de Vigilancia Epidemiológica. Ministerio de Salud Pública del Ecuador. 2014. Available: https://aplicaciones.msp.gob.ec/salud/archivosdigitales/documentosDirecciones/dnn/archivos/EDITOGRANNORMASIVE.pdf

23. Ross J, Larco D, Colon O, Coalson J, Gaus D, Taylor K, et al. Evolución de la resistencia a los antibióticos en una zona rural de Ecuador. Práctica Familiar Rural. 2020;5: 29–39. 10.23936/pfr.v5i1.144

24. Troya C, Herrera D, Guevara A, Obregón M, Gaus D, Larcos D, et al. Monitoreo local de Resistencia a los antibióticos en Escherichia coli en una zona rural de Ecuador: más allá del modelo biomédico. Práctica Familiar Rural. 2016;1. doi:10.23936/pfr.v1i1.128

25. OPS. Es fundamental la implementación de Programas de Optimización de Antimicrobianos (PROA). In: Organización Panamericana de la Salud [Internet]. 2021. Available: https://www.paho.org/es/noticias/17-11-2021-es-fundamental-implementacion-programas-optimizacion-antimicrobianos-proa

26. Edición Médica. Ecuador participa en proyecto latinoamericano de optimización de antimicrobianos en la UCI. Edición Médica. 2019. Available: https://www.edicionmedica.ec/secciones/profesionales/ecuador-participa-en-proyecto-latinoamericano-de-optimizacion-de-antimicrobianos-en-pacientes-de-uci--94737

27. Fabre V, Cosgrove SE, Secaira C, Tapia Torrez JC, Lessa FC, Patel TS, et al. Antimicrobial stewardship in Latin America: Past, present, and future. Antimicrobial Stewardship & Healthcare Epidemiology. 2022;2: e68. doi:10.1017/ash.2022.47

28. PAHO. Recommendations for Implementing Antimicrobial Stewardship Programs in Latin America and the Caribbean: Manual for Public Health Decision-Makers. Organización Panamericana de la Salud; 2018. doi:10.37774/9789275120408

29. PAHO. Latin American and Caribbean Network for Antimicrobial Resistance Surveillance (ReLAVRA+). In: DC Washington. 2023.

